# Regional and Sex-Specific Influences on BMI and Catheter Ablation in Paroxysmal Atrial Fibrillation

**DOI:** 10.1101/2025.10.24.25338737

**Authors:** Jay Nemade, Teimurazi Tchikohria, Gulnaz Dusekeeva, Arjun J Prakash, Nino Javakhidze, Roin Rekvava

**Author notes:** **<u>Corresponding Author:</u> Jay Nemade**, Faculty of Medicine, Georgian National, University SEU, 9 Tsinandali Street, Georgia, 0144, Georgia, **Email:**. **Funding:** No external funding was received for this study.

## Abstract

**Background:** Paroxysmal atrial fibrillation (PAF) is an intermittent arrhythmia with self-terminating episodes lasting fewer than seven days. Its development involves multiple factors, with body mass index (BMI) as a key modifiable risk factor. Elevated BMI promotes atrial remodeling and inflammation, increasing PAF susceptibility. Catheter ablation is now central to rhythm control and symptom management.

**Objective:** To evaluate regional and sex-specific patterns in BMI and catheter ablation selection among patients with PAF.

**Methods:** 84 PAF patients from Georgia (n = 43) and Kazakhstan (n = 41) were retrospectively analyzed. Age, sex, BMI, and ablation type were collected and stratified by center and sex. Inter-sex differences were assessed with t-tests, effect sizes calculated using Cohen’s d/ h, and BMI distributions evaluated using kernel density estimation.

**Results:** Analyses captured 97% of BMI variation. Females were generally older than males (d = 0.55), particularly in Kazakhstan (d = 0.70), while BMI differences were small and not statistically significant. Cryoablation was preferred overall (16% higher), with the largest sex disparity in Kazakhstan (42.9% higher in males); Georgia showed slight RFA preference in females (8.3%).

**Conclusion:** This study of PAF patients in Kazakhstan and Georgia reveals regional- and sex-specific variations in age, BMI, and ablation selection. We highlight the need for tailored risk assessment, early interventions, and awareness of potential clinical biases to ensure equitable care.

## Introduction

### Background

Atrial fibrillation (AF) is the most common cardiac arrhythmia, defined by disorganized atrial electrical activity that results in an irregular and often rapid ventricular response. It significantly increases the risk of severe complications such as stroke, systemic embolism, heart failure, and dementia, making it a leading cause of cardiovascular morbidity and mortality worldwide. [1]

AF pathogenesis is multifactorial, influenced by interrelated risk factors and comorbidities like hypertension, psychological stress, diabetes mellitus, and coronary artery disease. While these risk factors contribute to the development of AF, body mass index (BMI) stands out as a critical risk determinant due to its direct impact on clinical outcomes and its alterability with lifestyle interventions. [2–4]

### Role of BMI in AF

Elevated BMI, particularly in the obese range, promotes structural and electrical remodeling of the atria, largely through increased epicardial adipose tissue (EAT). EAT disrupts conduction and releases pro-inflammatory cytokines and adipokines, promoting atrial fibrosis and facilitating AF onset and maintenance. [5]

Obesity also exacerbates comorbidities such as arterial hypertension, diabetes mellitus, and ischemic heart disease (IHD) [6]. These comorbidities independently contribute to atrial remodeling and fibrosis. Thus, they create a synergistic environment that increases the risk of AF onset and progression [7]. Maintaining a balanced BMI is essential, because being underweight is also associated with higher all-cause and cardiovascular mortality in AF patients [8].

### Paroxysmal AF and it’s Management

Paroxysmal atrial fibrillation (PAF), a subtype of AF is characterized by episodes that start and terminate spontaneously. PAF episodes typically last from a few minutes up to seven days, without the need for medical intervention. [9]

PAF management includes pharmacological and non-pharmacological strategies to control symptoms and prevent progression. Catheter ablation both cryoablation & radiofrequency ablation (RFA), target pulmonary vein triggers. Ablation is increasingly used in symptomatic or drug-refractory patients to improve rhythm control and quality of life. [10]

### Research Gap

Understanding BMI distributions and inter-sex differences facilitates early risk stratification in PAF patients. Further, this allows targeted interventions for preventing progression to persistent / permanent forms.

While prior studies [11–15] have examined BMI and sex trends in AF, few [16–20] have focused specifically on PAF, and even fewer have investigated these patterns in the context of catheter ablation outcomes across different populations. Furthermore, variations in lifestyle, clinical context, and measurement timing can influence BMI distributions, highlighting the need for population-specific analyses.

## Methodology

### Study population & Data collection

A total of 84 patients were retrospectively included in this cross-sectional study, comprising 43 from Georgia (2022–2024) and 41 from Kazakhstan (2019–2022). To examine inter-sex differences in demographics, management strategies, and BMI among patients with PAF, data on age, BMI, sex, and procedure type were collected.

The two cohorts were collected over different time frames to reflect the periods during which standardized PAF management protocols were consistently implemented at each center. This approach allows for comparison of inter-sex BMI patterns while accounting for procedural standardization within each center.

### Statistical Analysis

Age, BMI, and procedure type were analyzed across the study cohort. Data from each center were stratified by sex, resulting in four subgroups: Georgia (male, female) and Kazakhstan (male, female). This stratification enabled both pooled and center-specific analyses while accounting for sex-related differences.

Preliminary analyses were conducted in the overall cohort as well as within individual strata. The normality of continuous variables was assessed using the Shapiro–Wilk test. Effect sizes were calculated to quantify differences: Cohen’s *d* for inter-sex differences in age and BMI, and Cohen’s *h* for differences in procedure type. Independent t-tests were used to evaluate sex differences between different strata. Given the moderate sample size, probability density functions (PDFs) for BMI were estimated using kernel density estimation (KDE) to evaluate the distributional characteristics and potential outliers.

All statistical analyses were conducted using Python 3.11.1 with the NumPy, SciPy, pandas, and scikit-learn libraries. Visualizations were generated using Bokeh. Two-tailed p-values <0.05 were considered statistically significant.

### Ethics

This retrospective study was conducted in accordance with local regulations, institutional guidelines, and the principles of the Declaration of Helsinki. Formal ethics committee approval was waived by the Personal Data Protection Service of Georgia, as the study involved only de-identified, routinely collected data. All patient information was handled confidentially to ensure privacy and compliance with ethical standards.

## Results

### Preliminary Data Analysis

From the baseline characteristics (Table 1) in the overall cohort, mean age of the patients was 62.9 ± 9.8 years (Mdn 65, range 29–80) and mean BMI was 30 ± 5.2 kg/m^2^ (Mdn 29.6, range 19.1–43.7). Cryoablation was performed in 58.3% and RFA in 41.7% patients. Males had mean age 60.2 ± 9.8 years and BMI 29.3 ± 4.6 kg/m^2^, with 62.5% undergoing cryoablation. Females had mean age 65.4 ± 9.3 years and BMI 30.8 ± 5.7 kg/m^2^, with 54.5% receiving cryoablation.

**Table 1.** Baseline Characteristics of the Cohort: Values are presented as mean ± standard deviation, median (IQR), and minimum–maximum for continuous variables. Categorical variables are shown as n (%).

| Group | Age (Years) | BMI (kg/m<sup>2</sup>) | Cryoablation | RFA |
| --- | --- | --- | --- | --- |
| **Complete (n = 84)** | 62.9 ± 9.8; 65 (57–68);<br>29–80 | 30 ± 5.2; 29.6 (26.1–33.7); 19.1–<br>43.69 | 49 (58.3%) | 35 (41.7%) |
| Male (n = 40) | 60.2 ± 9.8; 61 (57.5–66);<br>43–76 | 29.3 ± 4.6; 29.1 (26.1–32.7); 22–<br>43.7 | 25 (62.5%) | 15 (37.5%) |
| Female (n = 44) | 65.4 ± 9.3; 67 (58.3–71);<br>29–80 | 30.8 ± 5.7; 30 (26.2–33.7); 19.1–<br>43 | 24 (54.5%) | 20 (45.5%) |
| **Kazakhstan (n = 41)** | 60.6 ± 8.6; 61 (56–65);<br>37–77 | 29.4 ± 4; 29.4 (26.3–32.8); 21.6–<br>37.2 | 28 (68.3%) | 13 (31.7%) |
| Male (n = 21) | 57.8 ± 9.7; 60 (51–65);<br>37–73 | 28.6 ± 3.8; 28 (25.3–31.2); 23.2–<br>35.5 | 15 (71.4%) | 6 (28.6%) |
| Female (n = 20) | 63.5 ± 6.3; 64 (56–68);<br>53–77 | 30.3 ± 4; 31.3 (27.2–33.7); 21.6–<br>37.2 | 13 (65%) | 7 (35%) |
| **Georgia (n = 43)** | 65 ± 10.4; 67 (60–74); 29–<br>80 | 30.7 ± 6.2; 30.9 (25.5–36.3);<br>19.1–43.7 | 21 (48.8%) | 22 (51.2%) |
| Male (n = 19) | 62.8 ± 9.5; 66 (60–68);<br>40–76 | 30.1 ± 5.4; 29.5 (26–33.7);<br>22.1–43.7 | 10 (52.6%) | 9 (47.4%) |
| Female (n = 24) | 60.9 ± 11; 69 (63.3–74);<br>29–80 | 31.2 ± 6.8; 30.9 (25.5–36.3);<br>19.1–43 | 11 (45.8%) | 13 (54.2%) |

In Kazakh patients, mean age was 60.6 ± 8.6 years and BMI 29.4 ± 4 kg/m^2^. Cryoablation was performed in 68.3% and RFA in 31.7%. Males had mean age 57.8 ± 9.7 years and BMI 28.6 ± 3.8 kg/m^2^, with 71.4% undergoing cryoablation. Female had mean age 63.5 ± 6.3 years and BMI 30.3 ± 4 kg/m^2^, with 65% receiving cryoablation.

In Georgian patients, mean age was 65 ± 10.4 years and BMI 30.7 ± 6.2 kg/m^2^. RFA was slightly more frequent (51.2%) than cryoablation (48.8%). Males had mean age 62.8 ± 9.5 years and BMI 30.1 ± 5.4 kg/m^2^, with 52.6% undergoing cryoablation. Females had mean age 60.9 ± 11 years and BMI 31.2 ± 6.8 kg/m^2^, with 45.8% receiving cryoablation.

### Statistical Analysis

Normality of BMI distribution was confirmed across all groups using the Shapiro–Wilk test (all *p* > 0.05): complete cohort (*p* = 0.444 overall; 0.876 females; 0.101 males), Kazakhstan cohort (*p* = 0.545 overall; 0.497 females; 0.072 males), and Georgia cohort (*p* = 0.605 overall; 0.709 females; 0.588 males).

Normality testing for age indicated mixed distributions across cohorts. In the complete dataset, age was non-normally distributed overall (*p* = 0.002) and among females (*p* = 0.001), but approximately normal among males (*p* = 0.056). Within cohorts, age distribution in Kazakhstan was normal (*p* = 0.407 overall; 0.840 females; 0.389 males), whereas in Georgia, normality was not supported (*p* < 0.05 across all groups).

For BMI, the effect size was small overall (d = 0.27) and in Georgia (d = 0.17), but moderate in Kazakhstan (d = 0.42), reflecting a larger female–male difference there. For age, effect sizes were moderate to large overall (d = 0.55) and in Kazakhstan (d = 0.70), and moderate in Georgia (d = 0.40), indicating the strongest sex differences in Kazakhstan.

Between-sex differences in procedural management were small. For cryoablation, Cohen’s *h* was −0.16 overall, −0.14 in Kazakhstan, and −0.14 in Georgia, indicating slightly higher male uptake. For RFA, Cohen’s *h* was 0.16 overall, 0.14 in Kazakhstan, and 0.14 in Georgia, reflecting the reciprocal pattern.

Between-sex comparisons using independent-samples t-tests showed no significant differences in BMI overall (t = 1.26, p = 0.21), in Kazakhstan (t = 1.34, p = 0.19), or in Georgia (t = 0.56, p = 0.58). Age differences were significant overall (t = 2.50, p = 0.014) and in Kazakhstan (t = 2.25, p = 0.031), but not in Georgia (t = 1.32, p = 0.19).

### Distribution Analysis

KDE (Figure 1), consistent with the statistical tests, indicated that BMI distributions were approximately normal across the cohort having an area under curve (AUC) of 0.97 for the interval in the real data ranges. The overall BMI distribution had a slight positive skewness (γ_1_ = 0.35), and slightly platykurtic shape (γ_2_ = −0.22). Male BMI distribution was moderately right-skewed (γ_1_ = 0.78) and somewhat peaked (γ_2_ = 0.61), whereas female BMI distribution was nearly symmetric (γ_1_ = 0.02) with a slightly flattened peak (γ_2_ = −0.52).

**Figure 1.**
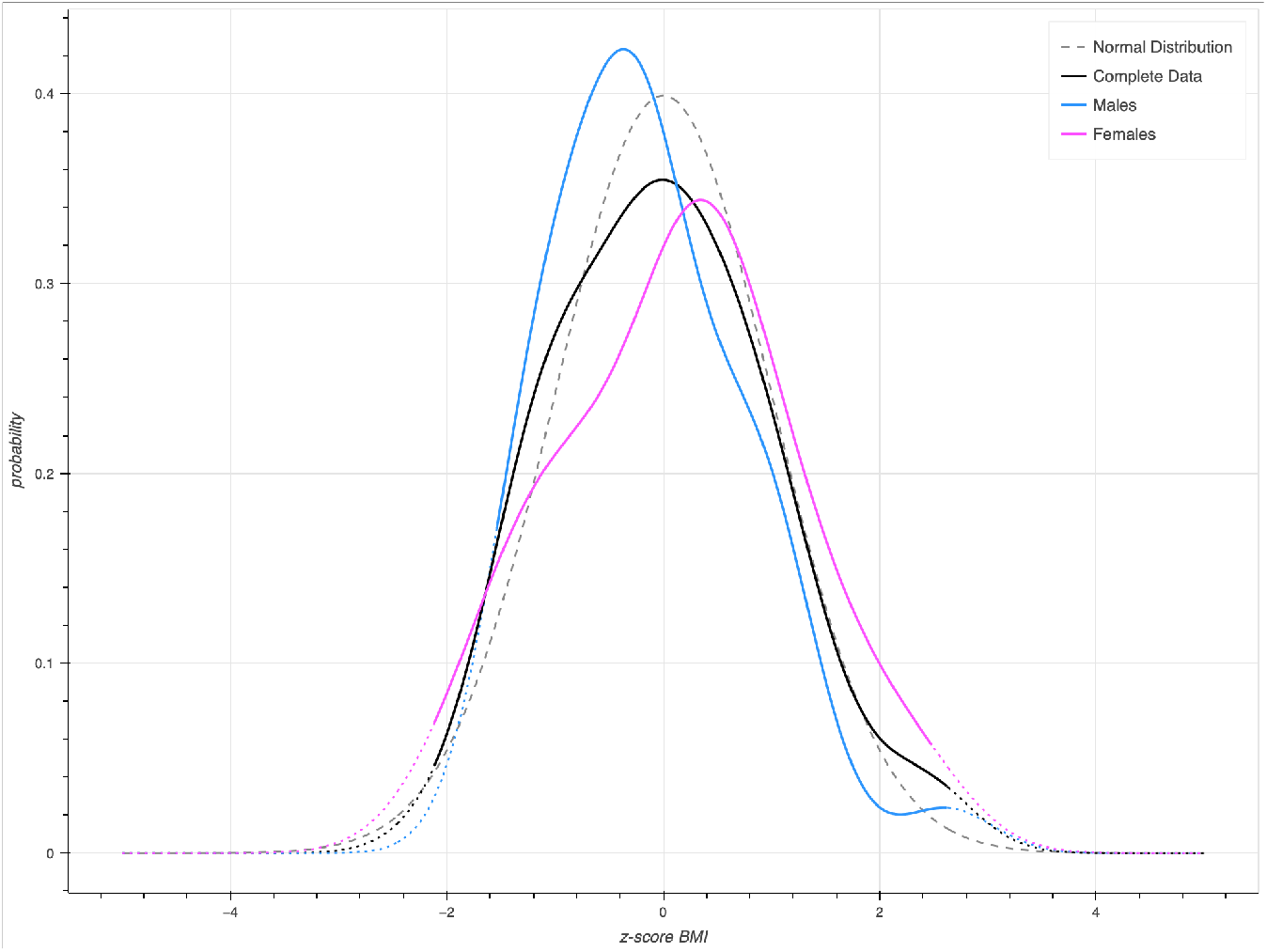
Kernel density estimation (KDE) of BMI Probability Density Functions (PDFs): The graph shows z-score vs probability of BMI, where the dotted lines show extended data, and the bold lines show actual data range of our cohort. A normal distribution is also plotted for reference.

## Discussion

### Principal Findings & Clinical Takeaways

The overall BMI distribution exhibited slight platykurtosis, implying relatively fewer extreme values. Additionally, despite a modest sample size, we capture 97% of BMI variation in our cohort. Thus, cautious extrapolation to Kazakh and Georgian populations with similar profiles is justified.

Females were generally older than males, most notably in Kazakhstan (Cohen’s d = 0.70; t = 2.25, p = 0.031), whereas Georgia showed a small, non-significant age difference favoring males (d = 0.40; t = 1.32, p = 0.19). Thus, regional variability may influence patient selection and timing of interventions.

Consistent with the opposite skewness, although females exhibited slightly higher BMI than males (30.8 vs. 29.1 kg/m^2^; d = 0.27), differences were not statistically significant overall or regionally. The inter-sex BMI gap was larger in Kazakhstan (1.7 kg/m^2^) than Georgia (1.1 kg/m^2^), suggesting local lifestyle or environmental contributions. While these trends are statistically non-significant, their consistency with mechanistic rationale permit insight into risk profiles and may inform preventive strategies.

Cryoablation was generally preferred over RFA. The greatest sex disparity was observed in Kazakhstan among males (42.9%). Georgia showed a slight RFA preference in females (8.3%). These patterns indicate potential sex- and region-specific biases in treatment selection. Therefore, clinician awareness to support equitable, evidence-based care is established.

### Future Directions

Additional variables such as socioeconomic status, comorbidities, and physician decision-making are needed to validate these findings. Evaluating long-term outcomes following cryoablation versus RFA across patient subgroups would further guide evidence-based, equitable interventions and inform regionally tailored clinical guidelines.

### Strengths and Limitations

Strengths include capturing nearly all BMI variation in a high-risk population and examining two distinct regional cohorts. Combined with our rigorous statistics, we effectively capture region- and sex-specific patterns.

Limitations involve the modest sample size and observational design, which restrict causal inference. Unmeasured factors, including comorbidities or physician preference, may have influenced outcomes. Furthermore, regional findings may not extend to other populations or healthcare settings.

### Conclusion

This is the first study to characterize regional and sex-specific BMI and procedural patterns in PAF patients from Kazakhstan and Georgia. We provide insights into demographic influences on catheter ablation selection.

Regional- and sex-specific variations in age, BMI, and ablation selection underscore the need for age- and sex-specific risk assessment. Our trends may inform sex specific early lifestyle interventions and risk stratification. We highlight potential clinical biases that warrant awareness to ensure equitable care. These findings provide novel insights on PAF management, informing risk stratification, procedural planning, and equitable care strategies in under-represented populations.

## Data Availability

All data produced in the present study are available upon reasonable request to the authors

## List of Abbreviations

BMI: Body Mass Index
EAT: Epicardial Adipose Tissue
AF: Atrial Fibrillation
PAF: Paroxysmal Atrial Fibrillation
RFA: Radiofrequency Ablation
SD: Standard Deviation
KDE: Kernel Density Estimation

## Notes

**Conflicts of Interest:** “None declared”

### Competing Interest Statement

The authors have declared no competing interest.

### Funding Statement

This study did not receive any funding

### Author Declarations

Formal ethics committee approval was waived by the Personal Data Protection Service of Georgia as the study involved only de-identified, routinely collected data. All patient information was handled confidentially to ensure privacy and compliance with ethical standards.

